# CT ECV Mapper: an interactive 3D Slicer application with a batch-capable pipeline for voxelwise CT-derived extracellular volume mapping of the liver and hepatic tumors

**DOI:** 10.64898/2026.08.09.26360018

**Authors:** Munemura Suzuki

## Abstract

**Background:** Extracellular volume fraction (ECV) derived from contrast-enhanced CT is a validated marker of hepatic fibrosis and has been reported to differ between hepatocellular carcinoma (HCC) and intrahepatic cholangiocarcinoma. In published work it is obtained from a small number of hand-placed two-dimensional regions of interest, and the software that computes it is either tied to one manufacturer’s workstation or based on spectral or dual-energy acquisition. We are not aware of an accessible tool that produces voxelwise liver ECV maps from conventional single-energy multiphase CT.

**Methods:** We developed CT ECV Mapper, a scripted 3D Slicer extension with a three-layer architecture whose numerical core imports neither slicer nor vtk and is unit-tested outside 3D Slicer. The interactive application provides two-stage registration that the operator inspects and accepts before any ECV is computed, operator-placed three-dimensional regions of interest, user-adjustable calculation parameters, a voxelwise ECV color map and ROI statistics; the same logic layer can be driven unattended across a cohort. The tool was applied to the 164 patients of the public WAW-TACE multiphase HCC/TACE dataset that have both unenhanced and delayed-phase series.

**Results:** 156 of 164 cases (95.1%) completed unattended. Whole-liver ECV had a median of 36.2% (interquartile range 31.9–41.5), consistent with published CT-ECV values for fibrotic and cirrhotic liver. Registering the arterial and portal phases on demand extended tumor ECV from the 38 lesions a conventional two-phase pipeline can reach to 248 lesions in 156 patients. Every failure was attributable to an identifiable mechanism: craniocaudal field-of-view mismatch between phases in six cases, aortic calcification within the blood-pool region in one, and in one case a labeling error in the source dataset, in which the series declared as unenhanced proved to be a second reconstruction of the portal venous phase — detected by the blood-pool validity check rather than by visual review.

**Conclusions:** Voxelwise CT ECV mapping of the liver and of hepatic tumors is feasible from conventional multiphase CT on an open platform, both interactively and as an unattended batch, with quality-control instrumentation that fails explicitly and diagnosably. This is a technical development and feasibility report; the application has not been evaluated against a reference standard and no claim of clinical validity is made.

## 1. Introduction

Hepatocellular carcinoma (HCC) is the most common primary liver cancer and arises, in the large majority of cases, on a background of chronic liver disease with progressive fibrosis or cirrhosis [1]. Transarterial chemoembolization (TACE) is the standard first-line locoregional treatment for intermediate-stage (BCLC-B) disease and is also used outside that stage when curative options are unavailable [2]. Patients worked up for TACE routinely undergo multiphase contrast-enhanced CT covering unenhanced, late arterial, portal venous and delayed phases — an acquisition that already contains, with no additional scanning and no additional contrast dose, the two phases required to compute the extracellular volume fraction (ECV).

ECV estimates the interstitial-plus-vascular compartment of a tissue from the equilibrium-phase partition of iodinated contrast between tissue and blood, corrected for hematocrit. In the liver, CT-derived ECV has been validated against biopsy-derived collagen-proportionate area [4] and correlates with histologic fibrosis stage [5] and with Child-Pugh class [6]; tumor ECV has been reported to differ between HCC and intrahepatic cholangiocarcinoma (ICC), plausibly reflecting differences in fibrous stromal content [7,8]. In essentially all of this literature, however, ECV is obtained by placing a small number of two-dimensional regions of interest (ROIs) by hand on the liver and the aorta and reporting a single scalar value per patient. That approach is operator-dependent, discards all spatial information about how ECV is distributed within an organ or within a lesion, and does not scale to cohort-sized analysis.

Cardiac MRI illustrates what the alternative looks like in mature form. Myocardial ECV mapping there is voxelwise, motion-corrected and fully automated, generated inline on the scanner console and presented to the reader as an image rather than as a hand-placed ROI measurement [9]. The clinical unit of analysis is a map. No comparable capability is in routine use for liver CT.

What is available for CT liver ECV today falls into two groups, neither of which closes that gap. Vendor workstation packages can compute ECV — Philips IntelliSpace Portal’s Multiphase Analysis and the dual-energy tools in Siemens syngo.via both appear in the published liver ECV literature — but they tie the analysis to one manufacturer’s workstation, and the studies using them are predominantly based on spectral or dual-energy CT, hardware that remains far from universally available. On the open-source side, we are not aware of an existing tool that produces voxelwise liver ECV maps from conventional single-energy multiphase CT; the one publicly available 3D Slicer ECV extension we identified targets cardiac MRI T1 mapping rather than CT. Neither group offers unattended batch processing across a cohort, which is what makes retrospective evaluation on existing data practical.

We therefore developed CT ECV Mapper, a scripted extension for 3D Slicer [11] that produces voxelwise liver and tumor ECV maps from conventional single-energy multiphase CT. The application is built around an interactive workflow — phase selection; a two-stage registration the user inspects and accepts before any ECV is computed; three-dimensional ROI definition for the blood pool, the target lesion and free-form measurement regions; and directly adjustable calculation parameters (assumed hematocrit, smoothing, optional median filtering, display range) that can be changed and recomputed without leaving the module — while its numerical core and logic layer remain independently scriptable, so that the identical computation can be driven unattended across an entire cohort.

This report describes that application and evaluates it on the publicly available WAW-TACE cohort [10]. The aims were: (1) to describe the design and functionality of the tool; (2) to demonstrate that the same code path runs unattended across a full cohort, and to characterize rather than suppress the technical failure modes and quality-control signals that emerge at that scale; and (3) to assess whether the resulting ECV values are plausible when set against published CT-ECV values and against general clinical expectation for this population. This is a technical development and feasibility report. It does not attempt to establish a new diagnostic threshold, and the physiological comparisons in Section 3 are made to test whether the tool produces credible output, not to advance a new biological claim.

## 2. Methods

### 2.1 Study population and data source

This is a retrospective secondary analysis of the publicly available WAW-TACE dataset (Zenodo record 12741586; CC-BY-4.0), comprising baseline four-phase (unenhanced/native, late arterial, portal venous, delayed) abdominal CT of 233 treatment-naïve patients with hepatocellular carcinoma (HCC) prior to transarterial chemoembolization (TACE), together with organ segmentations (TotalSegmentator), 377 hand-crafted tumor segmentations, and clinical metadata. Original data collection was approved by the Bioethics Committee at the Medical University of Warsaw (approval AKBE/41/2024); as a secondary analysis of a de-identified, publicly released dataset, no additional institutional review was sought.

Imaging protocol, per the original dataset publication: native phase, no contrast (N=200); late arterial phase, 15–30 s after contrast bolus (N=230); portal venous phase, 60–75 s (N=231); delayed phase, 4–5 min (N=193). Of the 233 patients, 164 had both native and delayed-phase series available and were included in the primary liver ECV pipeline.

The cohort was imaged on four different CT systems from four vendors (GE HealthCare Optima CT600, n=160; Siemens Healthcare Somatom Xceed, n=33; Philips Healthcare Ingenuity Core, n=20; Toshiba Medical Systems Aquilion One, n=20), with correspondingly heterogeneous acquisition parameters across the cohort (e.g. tube voltage 120 kVp in 214/233 examinations, 100–140 kVp in the remainder; median section thickness 1.5 mm for arterial/portal and 1.25 mm for delayed). As all four phases of a given patient are acquired in the same examination (and hence on the same scanner), this between-patient scanner heterogeneity does not by itself explain the within-patient, between-phase field-of-view differences characterized in this study (3.3); the cause of the latter was not identified and is reported as an open observation, not attributed to a specific mechanism.

### 2.2 Software architecture and interactive application

CT ECV Mapper was implemented as a scripted 3D Slicer (v5.8.0) extension with a three-layer architecture (Figure 2): (1) a Slicer-independent numerical core (ecv_core, pure NumPy/SciPy, unit-tested with pytest, n=74 tests) implementing the ECV formula, geometry utilities, and mask operations; (2) a Logic layer wrapping Slicer-specific registration and I/O calls; (3) an interactive GUI for single-case use. For this study, a batch driver script invoked the Logic layer directly across the full cohort, with per-case results appended incrementally to disk (JSON Lines and CSV) so that multi-hour batch runs could be safely interrupted and resumed without reprocessing completed cases.

The interactive application (Figure 1) presents the analysis as an ordered sequence of collapsible steps: selection of the unenhanced and delayed series; registration; definition of the blood-pool ROI; definition of a three-dimensional bounding box limiting the computation and display extent; computation of the ECV map; measurement within a two-dimensional circular ROI or a free-form three-dimensional ROI; and export. Registration is deliberately split into two separately triggered stages (2.3) so that the operator inspects and accepts each result before any ECV is computed, in keeping with the requirement that a registration be accepted before it is used quantitatively. The ECV map is rendered as a color overlay on the unenhanced series and can be reviewed against any other phase, or against another modality, in the same viewer.

**Figure 1.**
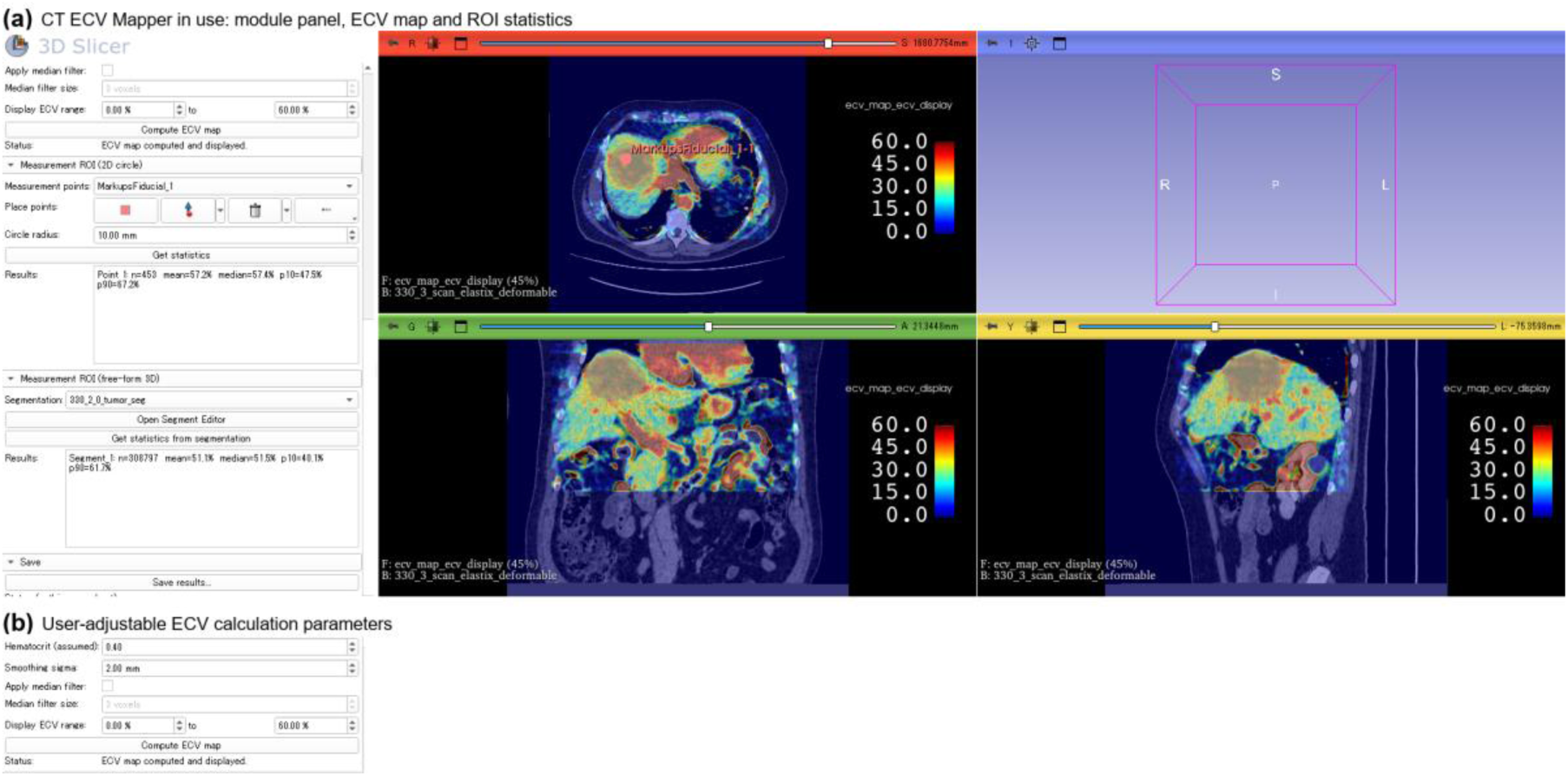
CT ECV Mapper in interactive use (case 330). (a) The module panel and the ECV map displayed over the unenhanced CT in axial, coronal and sagittal views, with the color scale in per cent. The lesion in the right hepatic lobe appears as a distinct high-ECV region against the background parenchyma. Statistics are shown for both measurement modes: a 2D circular ROI (10 mm radius; n = 453 voxels, mean 57.2%, median 57.4%) and a free-form 3D ROI (n = 308,797 voxels, mean 51.1%, median 51.5%), the latter using the tumor segmentation supplied with the dataset, loaded as a measurement ROI. (b) The user-adjustable calculation parameters. Smoothing sigma is 2.0 mm here for display clarity; the cohort analysis used the default 1.2 mm.

**Figure 2.**
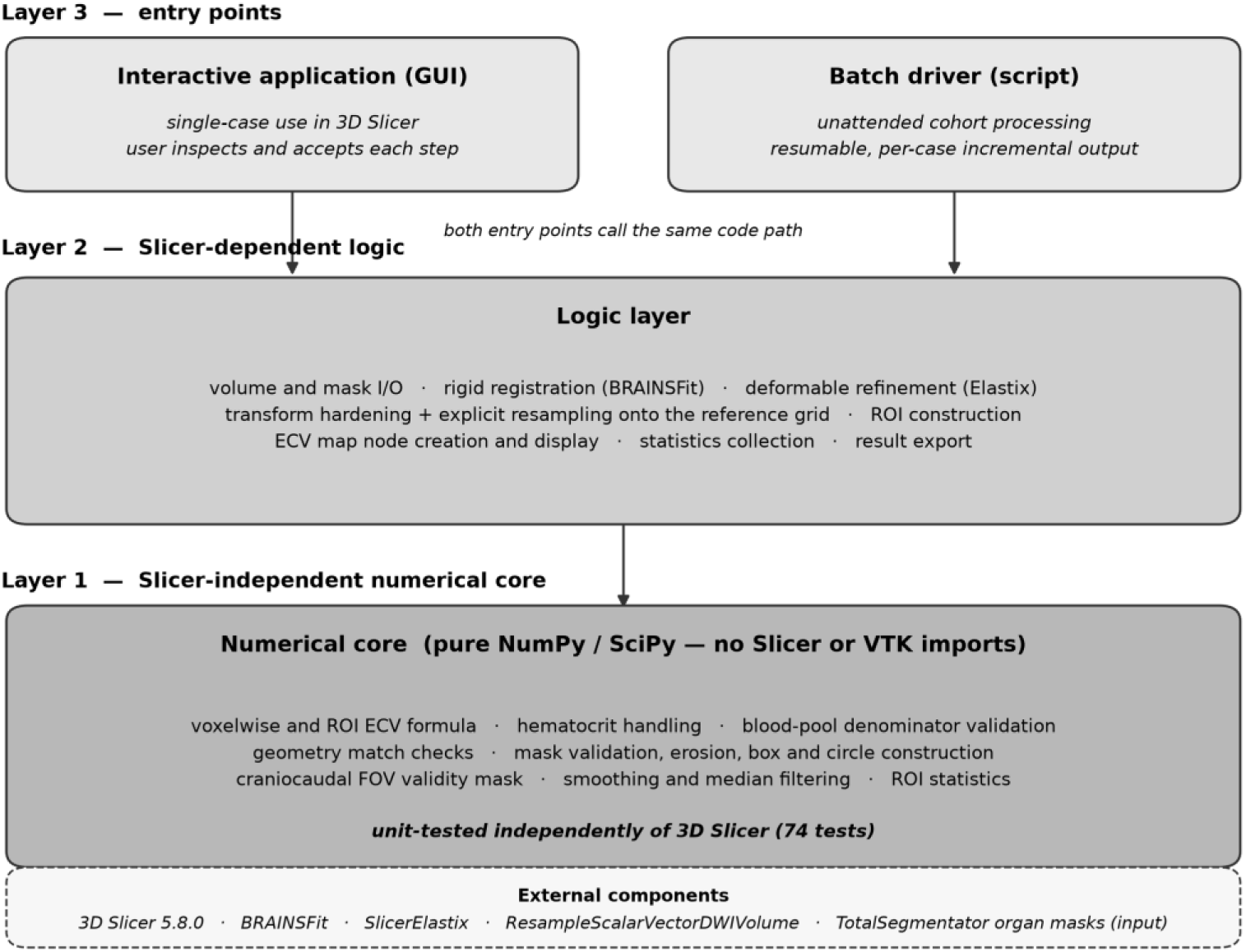
Three-layer software architecture. The interactive application and the batch driver are separate entry points that call the same Logic layer and the same Slicer-independent numerical core, so an interactive single-case result and a cohort batch result are produced by identical computation. The numerical core imports neither slicer nor vtk and is unit-tested outside 3D Slicer (74 tests).

Four quantities entering the calculation are exposed as editable fields rather than fixed constants: the assumed hematocrit (0.20–0.60, default 0.40), the Gaussian smoothing sigma applied identically to both phases before subtraction (0–5 mm, default 1.2 mm), an optional median filter (1–9 voxels), and the display range (default 0–60%). The sampling radius of the blood-pool ROI and of the circular measurement ROI are likewise user-set. Changing any of these and recomputing is a single action within the module, so the sensitivity of a given case to a given assumption can be examined directly rather than inferred. Only the display range is a pure visualization setting; the others change the computed values, and the values reported in Section 3 correspond to the defaults.

### 2.3 Image registration

Each non-native phase was registered to the native phase in two chained stages. First, rigid registration was performed with Slicer’s built-in BRAINSFit module (initializeTransformMode = “Off”). Second, deformable refinement was performed with Elastix (SlicerElastix extension, git revision 9ec57ae) using the unmodified “default0” preset, chained onto the BRAINSFit transform as the initial transform. This preset itself comprises two further sub-stages: an additional rigid refinement (EulerTransform) followed by a B-spline deformable stage (BSplineTransform, final control-point spacing 16 mm), both using advanced Mattes mutual information as the similarity metric and adaptive stochastic gradient descent as the optimizer, over 4 resolution levels (250 and 500 maximum iterations per level, respectively).

This preset was adopted as SlicerElastix’s own generic default and was not tuned for this application; its performance was judged only by qualitative visual review, by the author, a radiologist with over 20 years of clinical experience, on 2 representative cases prior to this study, not by a quantitative registration metric — stated here explicitly as a methods limitation.

Registration was originally implemented for the native–delayed pair only, and was extended in this study to register the native–arterial and native–portal pairs on demand, wherever a tumor segmentation or the dynamic multiphase analysis (2.9) required it. The exact parameter files are archived with the project source code for reproducibility.

### 2.4 Organ and tumor segmentation

Liver and aortic masks were taken from the dataset’s TotalSegmentator-derived organ segmentations (label 5 and 52, respectively; confirmed against TotalSegmentator’s official class map and by voxel-count spot check). The aortic mask was eroded by 2 mm (Euclidean distance transform) to reduce vessel-wall partial-volume contamination before blood-pool sampling.

Tumor masks were the dataset’s hand-crafted segmentations; each of the 377 tumor instances (233 patients; no patient had tumors segmented on more than one phase) was drawn on a single phase, heavily skewed toward the arterial (n=230) and portal (n=109) phases rather than native (n=4) or delayed (n=34). Each tumor mask was warped onto the native grid using the registration transform for its source phase (2.3), then explicitly resampled onto native’s exact grid (dimensions, spacing, origin, direction) via the ResampleScalarVectorDWIVolume CLI module with nearest-neighbor interpolation. This explicit resampling step was necessary because Slicer’s hardenTransform alone recomputes the output grid from the transformed volume’s bounding box rather than preserving the reference geometry, which had caused a shape mismatch in 16 of 17 tumor instances in an earlier iteration of this pipeline restricted to native/delayed-anchored masks.

The use of automated organ masks and of the dataset’s tumor segmentations throughout Section 3 was a consequence of requiring the cohort run to proceed unattended, and is not intrinsic to the method. In interactive use every ROI in the workflow is operator-defined (Figure 3): the blood-pool ROI is a point placed in the aorta with a user-set sampling radius rather than a derived vessel mask, and a measurement ROI may be drawn freehand in three dimensions, or placed as a circle, anywhere on the images — including directly on the computed ECV map — with the corresponding voxel statistics returned for that region. Any externally prepared segmentation can equally be loaded and used as a measurement ROI.

**Figure 3.**
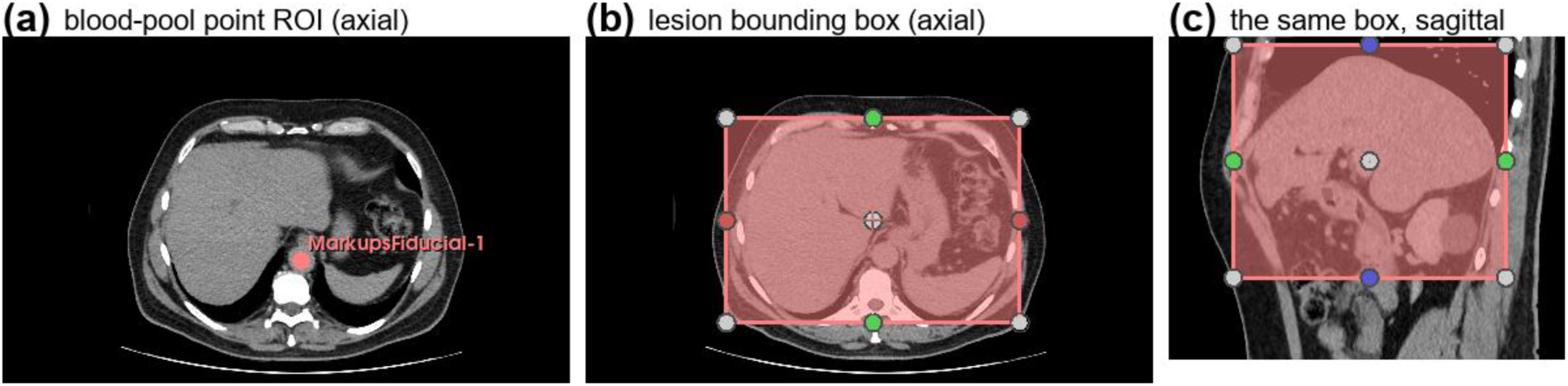
Operator-defined regions of interest (case 330). (a) Blood-pool ROI: a point placed by the operator in the aorta, with a user-set sampling radius; no vessel segmentation is involved. (b) Lesion bounding box in the axial plane and (c) the same box in the sagittal plane, positioned and resized by the operator, limiting the computation and display extent. Neither ROI requires an automated segmentation, and measurement ROIs are placed the same way (2.4).

### 2.5 Blood-pool sampling and hematocrit

Mean native-phase attenuation within the eroded aortic mask was used as the primary blood-pool baseline; the modal (most frequent, rounded to the nearest 1 HU) value was also computed as a measure less sensitive to focal non-calcified plaque/thrombus within the ROI.

This dataset provides no measured hematocrit. Per protocol, the primary analysis used a fixed assumed hematocrit of 0.40. As a secondary, exploratory comparison, a per-patient hematocrit estimate was derived from the modal native aortic attenuation via a formula reported for dual-energy CT virtual-unenhanced blood attenuation (Kim et al., AJR 2022; hematocrit [%] = 0.85 × HU − 5.40), clipped to [0.20, 0.60]. This formula was derived for a different acquisition type (dual-energy virtual-unenhanced) than this dataset’s true single-energy unenhanced phase and has not been separately validated; it is reported only as an exploratory secondary estimate, not a validated substitute for the fixed assumption.

### 2.6 Voxelwise ECV calculation

Native and delayed-phase volumes were Gaussian-smoothed (σ = 1.2 mm) after registration. Voxelwise ECV was computed as:

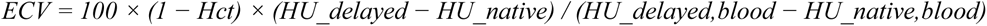

restricted to voxels with finite HU in [−500, 3000] and a minimum blood-pool enhancement of 1.0 HU (a case was marked failed if the whole-ROI mean blood enhancement itself did not clear this threshold). Raw ECV values were not clipped; only display ranges are clipped in the interactive GUI.

### 2.7 FOV-mismatch detection and exclusion

Preliminary review found the four phases of a given patient frequently differ in scan coverage (e.g. the delayed phase cropped to the upper abdomen) and, less often, in-plane field of view and slice thickness — properties not visible from the segmentation masks alone. To prevent resampler-fill/extrapolated voxels (present wherever the moving phase’s original coverage does not extend as far as native’s) from contaminating any ROI statistic, a geometry-only function (slice_fov_valid_mask) was implemented: for a given pair of phases, it determines which native-grid slices fall, by physical position, outside the moving phase’s original (pre-registration) scan range, using only the two volumes’ affine transforms (no dependency on Slicer APIs; unit-tested). Every liver, aortic, and tumor ROI statistic in this pipeline excludes voxels in slices flagged this way, per source phase.

### 2.8 Additional QC instrumentation

Beyond the pass/fail outcome of the pipeline itself, four supplementary per-case measures were recorded so that a case’s numerical result could be weighted by how much confidence it deserved, not treated as uniformly reliable simply because it completed:

i. FOV-exclusion extent — the fraction of the liver and aortic masks’ native-grid slices that 2.7 actually excluded for that case. This quantifies how much of the correction in 2.7 was invoked, distinguishing a case barely touched by it from one where a large fraction of the mask was affected.
ii. Residual flat-region (“plateau”) detector — within a given ROI, the fraction of voxels sharing the single most common rounded HU value. Real tissue attenuation, being continuous and noisy, essentially never repeats one exact value across more than a small fraction of an ROI; a large plateau fraction signals a suspiciously flat/constant region. Because this is computed AFTER the geometry-based exclusion in 2.7, a nonzero result here reflects a flat region 2.7 did NOT already remove — e.g. an in-plane (rather than craniocaudal) FOV mismatch, or a resampler artifact from some other cause — making it a complementary, model-free check rather than a duplicate of 2.7.
iii. Aortic attenuation sanity range — whether the mean native-phase aortic HU falls within a coarse physiological range for unenhanced blood (−30 to 150 HU). A value outside this range is a simple screen for gross contamination of the blood-pool ROI (e.g. dense calcification, a mislabeled phase, or a segmentation error) that would otherwise not be obvious from the ECV output alone.
iv. Liver valid fraction — the fraction of the anatomical liver mask that survived BOTH the FOV exclusion (2.7) and the HU-range/blood-enhancement validity filter (2.6) and therefore actually contributed to that case’s reported ECV statistics. A low value flags a case whose reported ECV, even though the pipeline did not fail outright, is based on a small and possibly unrepresentative fraction of the liver.

These four continuous measures were combined into a single summary qc_flags field per case using fixed thresholds, for convenience in filtering, but the underlying continuous values were retained throughout so that any threshold choice could be revisited during analysis rather than being baked in silently. In-plane FOV mismatch prevalence (3.3) was assessed separately, retrospectively, across the full 233-patient cohort directly from the NIfTI headers (in-plane extent and slice thickness of each phase versus native), independent of and without requiring registration — this is a cohort-level characterization, not a per-case flag applied by the pipeline itself.

### 2.9 Dynamic multiphase HU profile

As a hematocrit-independent complement to ECV, for every tumor-bearing case all four phases were registered to native (2.3), and mean HU was sampled, per phase, within the whole liver mask (once per case) and within each tumor mask, restricted to that phase’s own valid coverage (2.7). For each tumor instance, a washout index was defined as (peak − HU_delayed) / (peak − HU_native), where peak = max(HU_arterial, HU_portal); values near 1 indicate the tumor’s peak-over-baseline enhancement had returned to baseline by the delayed phase, values near 0 indicate no washout. A binary “washout sign” was also defined as HU_delayed(tumor) < HU_delayed(liver) for the same case (the CT correlate of the qualitative LI-RADS washout appearance).

### 2.10 Statistical analysis

Descriptive statistics are reported as mean ± SD and median [IQR] as appropriate. Two tumor instances confirmed to originate from identical source segmentation files (case 7) were deduplicated before any aggregate statistic. Associations between the washout index and ECV were assessed with Spearman’s rank correlation; the two washout-sign subgroups were compared with the Mann-Whitney U test. A sensitivity analysis recomputed descriptive ECV statistics excluding cases with any non-empty qc_flags (2.8) and two additional cases with an independently confirmed severe registration failure. All analyses were performed in Python (NumPy, SciPy, pandas); p<0.05 was considered significant, without correction for multiple comparisons given the exploratory nature of this analysis.

### 2.11 AI-assisted software development and analysis

The requirements, the three-layer architecture, the interactive workflow design, the data-handling rules, the choice of registration method, and the acceptance criteria for each processing step were specified by the author. Implementation of that specification — the extension source code, its unit test suite, the batch driver, the retrospective geometry-checking scripts, the statistical analyses and the figure-generation scripts reported here — was carried out with the assistance of a large language model (Claude, Anthropic) used as a coding and analysis assistant under the direction of the author. All study design decisions, all radiological interpretation, and the acceptance or rejection of every registration were made by the author. Analysis output was verified against the source data by the author, and errors introduced during AI-assisted development were identified and corrected in this way — including the organ-mask lookup defect described in 3.1, the transform-hardening defect described in 2.4, and a stale hardcoded statistic in an earlier version of Figure 5. The author takes full responsibility for the correctness of the code and of every value reported.

## 3. Results

### 3.1 Cohort and pipeline yield

Of 233 patients in the source dataset, 164 had both native and delayed-phase series and were processed by the liver ECV pipeline. Of these, 156 (95.1%) completed successfully and 8 (4.9%) failed the blood-pool enhancement validity check (2.6), after the two corrections described below (3.2, 3.3) (Table 1). No case in this cohort lacked a usable native-phase organ segmentation.

**Table 1.**
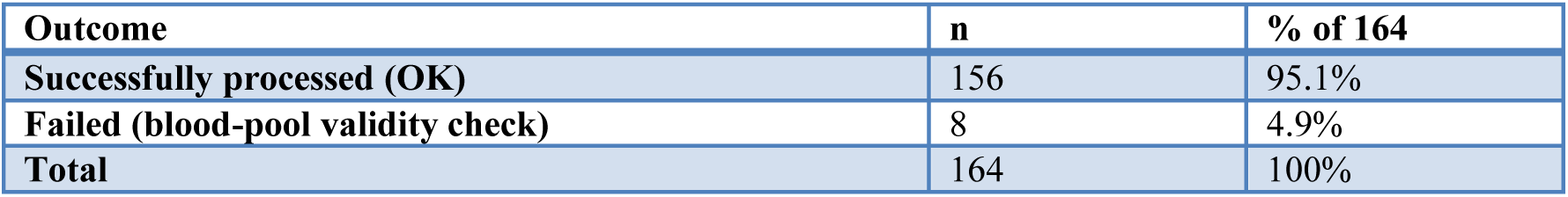
Liver ECV pipeline outcome, full cohort.

This last point required its own correction. An initial version of the pipeline looked up each case’s organ-mask archive member by reconstructing its expected filename, and treated a lookup failure as a missing segmentation; by that method, 49 of 164 cases (29.9%) appeared to lack a usable mask. Inspecting the archive directly found that 98 of its 854 filenames (11%) additionally embed the original DICOM series description (e.g. “…_total_segmentator.6_WATROBA_OLTX_20161028110238_2.nii.nii.gz”, “WATROBA” being Polish for liver) rather than following the plain pattern assumed by the lookup — and that all 49 “missing” cases in fact had a phase-0 mask present under one of these alternate names.

Indexing the whole archive up front instead of reconstructing filenames (2.4) resolved this: reprocessing the recovered 49 cases yielded 46 additional successes and 3 additional failures (2 by the craniocaudal FOV mechanism in 3.2, 1 by the blood-pool validity check), accounting for the difference between Table 1 and the organ-mask availability that would otherwise have been reported. This is stated explicitly because the original 29.9% figure would otherwise read as a limitation of the public dataset; it was not — the dataset’s own organ-mask coverage for this 164-case cohort is complete.

### 3.2 Effect of FOV-mismatch exclusion on pipeline yield

Before implementing the FOV-mismatch exclusion (2.7), 10 of 164 cases failed the blood-pool enhancement check, with the aortic ROI showing an implausible non-positive native-to-delayed attenuation change. A retrospective, registration-independent geometry check found the aortic mask’s craniocaudal extent fell outside the delayed phase’s original scan coverage by 30–526% in 9 of these 10 cases (a value >100% indicates the mask’s own extent barely or does not overlap the delayed phase’s coverage at all) — a mechanism the segmentation-derived organ mask itself does not reveal, and one that a liver-only version of the same check had missed in most of these cases (fixed at the source in 2.7, not merely flagged).

After implementing the exclusion, 5 of these 10 cases were fully rescued, with physiologically plausible positive blood-pool enhancement (27–45 HU) restored; the remaining FOV-affected cases instead failed explicitly (“aortic mask empty after excluding out-of-FOV slices”) rather than silently returning an implausible value. One case (101) remained failed with zero slices excluded by this mechanism. Review of the aortic ROI on the unenhanced series found frank calcification rather than soft plaque (12.8% of ROI voxels above 100 HU, maximum 512 HU, 95th percentile 181 HU), which inflates the unenhanced mean and so suppresses the native-to-delayed difference; the delayed phase itself was adequately enhanced in this case (liver 47.2 to 63.6 HU), and its unenhanced series is 5.0 mm thick against 2.5 mm for the other phases, so degraded sampling of a structure as narrow as the aorta plausibly contributes as well. This is a distinct failure mode not addressed by the field-of-view correction. After the organ-mask correction (3.1) made 49 further cases reachable, 2 of these failed by the same craniocaudal FOV mechanism (aortic mask entirely excluded) and 1 (case 369) by the blood-pool validity check, returning a negative aortic enhancement of −23.8 HU. This last case proved to be a labeling error in the source data rather than a processing failure, and is described separately below. In total, across the final cohort, 6 of 164 cases failed by the craniocaudal FOV mechanism, 1 of 164 by aortic calcification within the blood-pool ROI, and 1 of 164 because the series declared as unenhanced was not unenhanced (Table 2).

**Table 2.** Failure-mode taxonomy identified in this cohort.

| Failure mechanism | Cases | Status |
| --- | --- | --- |
| **Craniocaudal (z-axis) FOV mismatch** | 6 (158, 165, 539, 543, 33, 53) | Detected and corrected (2.7); these 6 fail explicitly rather than silently |
| **Aortic calcification within the blood-pool ROI** | 1 (101) | Characterized; not corrected. Coincident 2x section-thickness mismatch may contribute |
| **Source-data phase mislabeling (series declared unenhanced is contrast-enhanced)** | 1 (369) | Detected by the blood-pool validity check (2.6); a dataset error, not a pipeline error. Cohort audited, no other case affected |
| **In-plane (x/y) FOV / slice-thickness mismatch** | 30/233 cases (13%), cohort-wide | Characterized; not corrected |
| **Extreme inter-phase scan-length mismatch → registration non-convergence** | 1 (documented separately, not in Table 1's FAILED count) | Accepted as a data-driven limitation |
| **Organ-mask filename lookup (this pipeline's bug, see 3.1)** | 0 (all 49 recovered) | Fixed (2.4) |

Case 369 warrants its own description because the mechanism lies in the source data. Its declared unenhanced series returned an aortic attenuation of 93.9 HU and a liver attenuation of 78.7 HU, both far above any plausible unenhanced value, and its liver attenuation fell from that value to 66.4 HU on the delayed phase — impossible for a genuine unenhanced-to-equilibrium pair. Visual review by the author found the series declared as phase 0 and the portal venous series to be the same contrast phase, differing only in noise. Quantitatively the two agree to 0.9 HU in the liver and 0.5 HU in the aorta, cover the same craniocaudal extent (approximately 474 mm), and differ in reconstructed section thickness (1.25 versus 2.5 mm), consistent with two reconstructions of one portal venous acquisition of which the thinner was filed into the unenhanced slot. The blood-pool validity check (2.6) therefore did not merely fail safely on this case; it identified a mislabeled series in a public dataset that would otherwise have yielded a plausible-looking but meaningless ECV value.

Because that failure mode would invalidate any case it affected, the remaining cohort was audited for it. Across the 156 successfully processed cases, liver attenuation on the declared unenhanced series ranged from 23.6 to 67.7 HU (5th–95th percentile 31.4–56.5, median 45.0), with no case exceeding 70 HU; case 369’s 78.7 HU lies outside that range entirely. Among the eight failures, only case 369 shows the pattern. We therefore found no evidence that any case contributing to the ECV results reported here carries a comparable phase-labeling error. This audit compares declared labels against image content as a quality-control step; phase assignment itself was always taken from the dataset’s filenames and manifest, never inferred from attenuation.

### 3.3 In-plane FOV mismatch (documented, uncorrected)

Independent of registration, comparing each phase’s in-plane field of view and slice thickness to native’s across all 233 patients found 30 (12.9%) with an in-plane FOV difference exceeding 5% on at least one phase, 7 (3.0%) exceeding 20%, and slice thickness differing from native in 236 of 562 (42.0%) phase pairs. This axis of FOV mismatch is not addressed by the exclusion in 2.7 (which is craniocaudal only) and is reported here as a characterized but uncorrected limitation.

### 3.4 Liver ECV

Among the 156 successfully processed cases, whole-liver ECV (Hct=0.40) had a median of 36.2% (IQR 31.9–41.5). The arithmetic mean, 38.6% (SD 20.2), is inflated by three cases that returned physiologically impossible values (−13.8%, 143.1% and 232.9%, all three independently QC-flagged); excluding those it is 37.0%, while the median is unchanged. Median and interquartile range are therefore used as the primary summary here and throughout. This range is consistent with published CT-ECV values for cirrhotic liver parenchyma (e.g. Child-Pugh A/B/C: 29.7/35.6/45.3%; a separate cohort: 25.0/28.3/33.6% for F0-1/F2-3/cirrhosis), as expected for a TACE-eligible HCC cohort with a predominantly fibrotic/cirrhotic background liver. An example map with its four source phases is shown in Figure 4.

**Figure 4.**
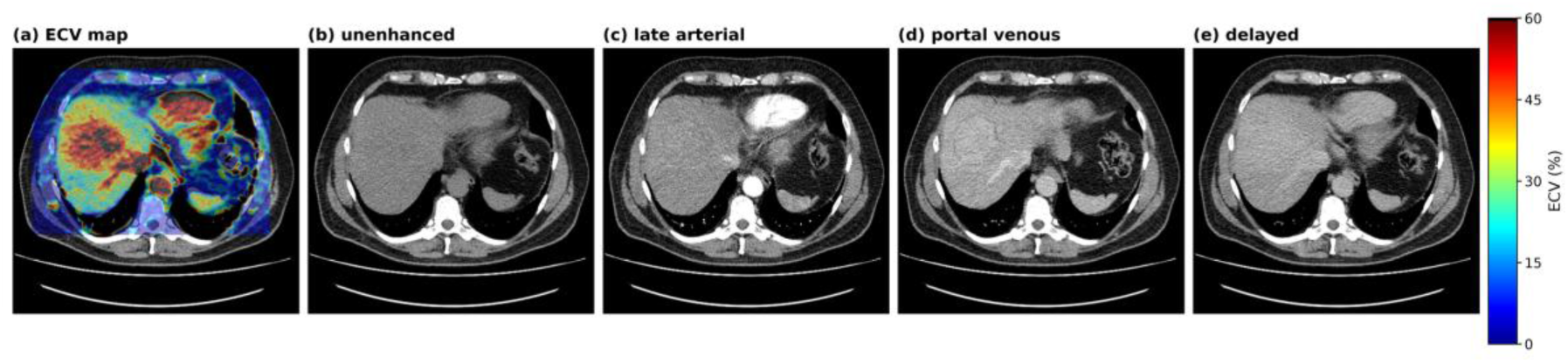
ECV map alongside its source phases at the same axial level (case 330, z = 1674.4 mm). (a) ECV map overlaid on the unenhanced CT at 45% opacity, with the scale in per cent; the map is computed only within the operator-placed bounding box (2.2, Figure 3), which is why the overlay ends at a rectangular boundary. (b–e) The four acquired phases, displayed at a common window (width 350 HU, level 40 HU). The map can be reviewed side by side with any phase, or with another modality, giving a spatially resolved result rather than a single value per patient. Panels are rendered directly from the volumes; the ECV map is the one exported by the application’s own save function, with the parameters recorded in Figure 1.

**Figure 5.**
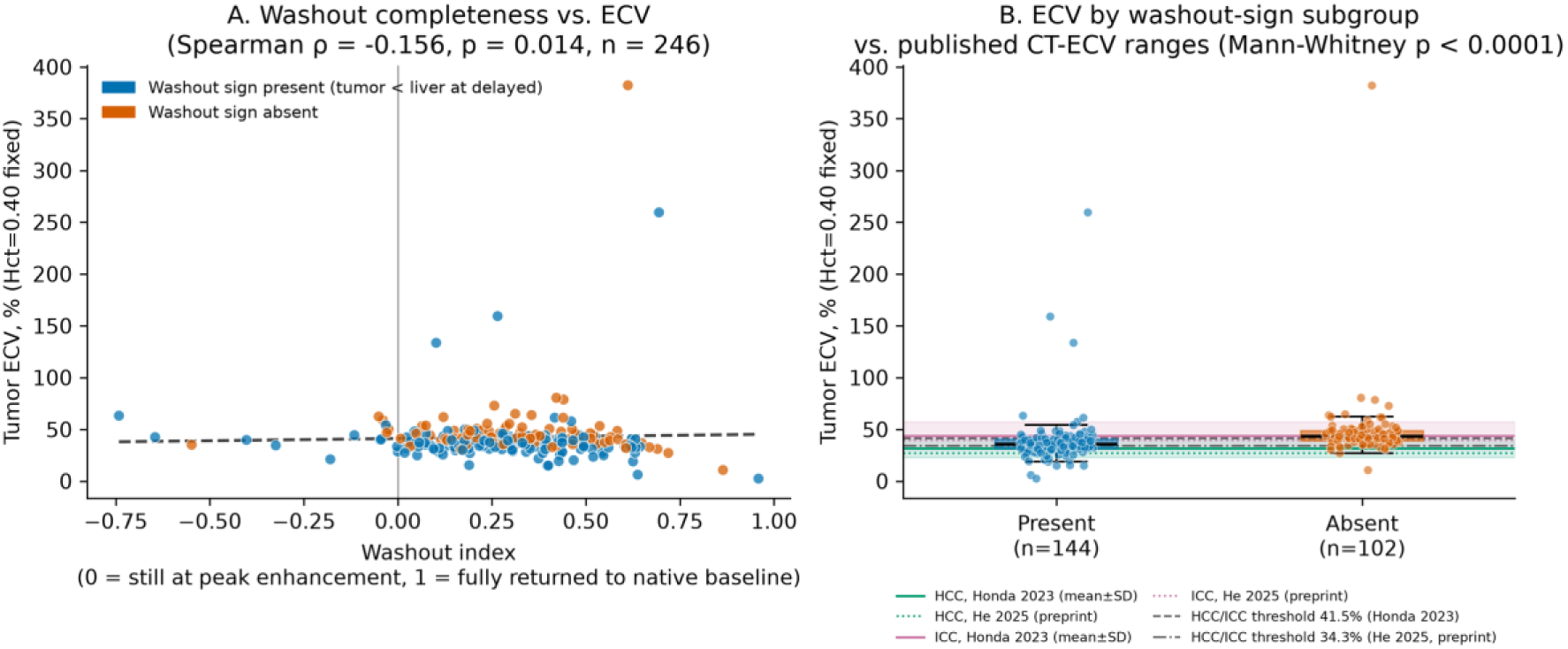
Hematocrit-independent consistency check (n = 246 tumor instances). (A) Washout index versus tumor ECV, with linear trend. The Spearman correlation shown does not survive adjustment for the delayed-phase tumor attenuation that both quantities contain (partial ρ = −0.04, p = 0.57); it is reported as an internal consistency check on the implementation, not as an independent association (4.5). (B) Tumor ECV by washout-sign subgroup, with published CT-ECV reference bands for HCC and intrahepatic cholangiocarcinoma (mean ± SD, Honda et al. 2023) and both published HCC/ICC discrimination thresholds (41.5%, Honda et al. 2023; 34.3%, He et al. 2025, preprint). Boxes show median and interquartile range with whiskers; individual instances are overplotted. All annotated statistics are computed from the plotted data.

### 3.5 Tumor ECV: pipeline extension and distribution

Restricting tumor ECV to native-or delayed-phase-anchored masks only (the pipeline’s original scope) would have covered 38 tumor instances in 33 patients. Extending registration to the arterial and portal phases (2.3–2.4) increased this to 248 tumor instances in 156 patients (100% technical success once a case’s liver processing succeeded), with tumor masks originating from the arterial (n=153), portal (n=65), delayed (n=27), and native (n=3) phases. Median tumor ECV (Hct=0.40) was 39.3% (IQR 33.6–44.8); the mean of 42.4% (SD 29.4) falls to 39.3% once four instances returning values above 100% are excluded, all four QC-flagged. The median is modestly higher than published CT-ECV means for HCC (26.9% [He et al., Research Square preprint 2025] to 31.6% [Honda et al., Front Oncol 2023]) and closer to published values for intrahepatic cholangiocarcinoma (39.7–43.8%, same two sources respectively); the two studies’ reported HCC/ICC discrimination thresholds were 34.3% (sensitivity 79.4%, specificity 83.3%; unreviewed preprint) and 41.5% (AUC 0.763; peer-reviewed), respectively.

### 3.6 Relationship between multiphase washout pattern and ECV

In the 246 evaluable tumor instances with complete 4-phase sampling (two duplicate instances removed, 2.10), the washout index (2.9) had a median of 0.29 [IQR 0.18–0.44], indicating that on average roughly 70% of peak enhancement remained at the delayed phase. A CT washout sign (2.9) was present in 144/246 (58.5%) and absent in 102/246 (41.5%). Washout index correlated negatively with ECV (Spearman ρ = −0.156, p = 0.0142).

ECV differed markedly between the two washout-sign subgroups: instances with a washout sign had a median ECV of 35.6% (mean 38.5%), closely matching the published HCC range, while instances without a washout sign had a median ECV of 43.5% (mean 47.9%), closer to the published ICC range (Mann-Whitney U, p<0.0001; Figure 5). A sensitivity analysis excluding all instances with a non-empty qc_flags value and two additional cases with an independently confirmed severe registration failure changed the overall tumor ECV median only marginally (39.0%, n=159), indicating the elevation relative to literature is not attributable to a small number of known-compromised cases.

The four-phase sampling itself was also audited, since the arterial and portal phases enter the washout index but are not protected by the craniocaudal exclusion in the way the unenhanced– delayed pair is. Four tumor instances returned a negative mean attenuation in at least one phase, which is impossible for soft tissue and indicates a mask lying partly outside the moving phase’s true coverage (in one instance all four phases were near −180 HU); two further cases returned a delayed-phase liver attenuation below 30 HU. Excluding all eight affected instances left the results of this section essentially unchanged (washout-sign-positive median ECV 35.6%, negative 43.6%, n = 238; Mann-Whitney p = 2.0 × 10⁻¹³; washout index median 0.287), so the comparison is not driven by them. It does, however, identify a gap: the dynamic sampling carries no plausibility filter of its own, and the negative values imply in-plane rather than craniocaudal mismatch, an axis the exclusion in 2.7 does not address.

### 3.7 Sensitivity to aortic-HU statistic and hematocrit assumption

Mean and modal native aortic attenuation diverged systematically (mean 43.8 ± 24.3 HU vs. mode 33.7 ± 9.8 HU across 156 cases; Spearman ρ = 0.644), with the largest divergences occurring in cases independently flagged for aortic plaque, thrombus, or an out-of-range aortic HU. Using the aorta-HU-derived hematocrit estimate (2.5; median estimated Hct 0.235, versus the fixed 0.40) in place of the fixed assumption raised both liver and tumor ECV by a median of approximately 9.9–10.2 percentage points, moving tumor ECV further from, not closer to, the published HCC range; this was taken as further support for reporting the fixed Hct=0.40 assumption as primary.

## 4. Discussion

### 4.1 Summary

We developed CT ECV Mapper, an interactive 3D Slicer application that generates voxelwise ECV maps of the liver and of hepatic tumors from conventional single-energy multiphase CT, with user-adjustable calculation parameters, three-dimensional ROI-based measurement, and a scriptable logic layer that allows the identical computation to be run unattended over a cohort.

Applied to 164 patients of the public WAW-TACE dataset, the batch path completed 156 cases (95.1%), extended tumor ECV coverage from the 38 instances a conventional two-phase pipeline could reach to 248 instances in 156 patients by registering the arterial and portal phases on demand, and produced liver and tumor ECV values consistent with published ranges. The eight failures were traced to two identifiable and reportable mechanisms rather than remaining unexplained.

### 4.2 Relation to existing tools

The gap this application addresses is one of access rather than of algorithm: the ECV calculation itself is long established, but the software that implements it for the liver is either bound to a specific vendor’s workstation or, in the published literature, replaced by manual ROI placement. Two consequences follow. First, because vendor implementations in the liver ECV literature are largely spectral or dual-energy, departments operating conventional single-energy scanners — the majority — have had no straightforward route to ECV at all; CT ECV Mapper deliberately targets that conventional acquisition. Second, a voxelwise map is a different measurement from a hand-placed ROI value, and this distinction turns out to matter for interpretation (4.4). In this respect the design goal was the one already realized in cardiac MRI, where automated pixelwise ECV maps are produced inline and read as images [9]; the contribution here is not a new formula but a usable, vendor-neutral implementation of an existing one, on an open platform, with a batch path attached.

Two properties of the CT route are worth stating explicitly, because they are what make this kind of retrospective work possible at all. First, no dedicated quantitative sequence is required: MR ECV depends on pre- and post-contrast T1 mapping acquired for that purpose, so it must be planned prospectively, whereas the two phases CT ECV needs are already present in any standard multiphase liver protocol — which is why a 164-patient cohort could be analyzed here from an archive assembled for an unrelated purpose. Second, CT attenuation is insensitive to flow, so the aortic blood-pool reference is not subject to the inflow effects that complicate blood-pool T1 measurement in MR; the denominator of the ECV calculation is correspondingly better behaved.

The output being a voxelwise map rather than a scalar also changes what can be asked of it. Because the map is registered to the unenhanced series, it can be displayed alongside any acquired phase or another modality (Figure 4), so whether ECV adds anything to visual interpretation becomes a question that can be examined directly rather than assumed. Spatial resolution likewise permits regional comparison against pathology, which a single per-patient value cannot support; the heterogeneity visible within the example lesion in Figure 1 is precisely the information a representative ROI discards.

### 4.3 Plausibility of liver ECV output

Whole-liver ECV had a median of 36.2% (IQR 31.9–41.5) across the 156 successfully processed cases. Published CT-ECV values for the fibrotic and cirrhotic liver span roughly 25–34% across fibrosis stages F0-1 to cirrhosis [5] and 29.7%, 35.6% and 45.3% for Child-Pugh A, B and C respectively [6]. A TACE-eligible HCC cohort is expected to sit in the cirrhotic part of that distribution, and the observed median does. We regard this as the tool’s most direct plausibility evidence, with one methodological qualification: both comparator series derive ECV from iodine maps on spectral or dual-energy CT, whereas this application computes it by attenuation subtraction on single-energy CT. Agreement of range across those two methods is supportive but is not a method-matched validation, and no such validation is claimed here. We note also that the arithmetic mean of this distribution (38.6%) is inflated by three cases returning physiologically impossible values (−13.8%, 143.1%, 232.9%); excluding them the mean is 37.0%, while the median is unchanged. All three were independently flagged by the quality-control instrumentation (4.6), and the median with interquartile range is the more appropriate summary throughout.

### 4.4 Plausibility of tumor ECV output, and why a map is not an ROI

Tumor ECV had a median of 39.3% (IQR 33.6–44.8) across 248 instances; the mean of 42.4% falls to 39.3% once four instances returning values above 100% are excluded, all four of which were QC-flagged. This is modestly higher than published CT-ECV means for HCC, reported as 26.9% [8] and 31.6% [7], and closer to published values for ICC (39.7% [8], 43.8% [7]) despite the source cohort containing only HCC. We do not read this as evidence about tumor biology, and specifically not as evidence of misclassification, since the cohort’s HCC diagnoses were established independently of this tool. The most likely explanation is that the two numbers are not the same measurand. Published tumor ECV is obtained from a small ROI placed by an operator on representative tumor tissue, typically on the few largest cross-sections; the value reported here is a whole-tumor volumetric mean over every voxel of an expert segmentation, which necessarily includes necrotic, hemorrhagic and peripherally partial-volume-averaged voxels that a human placing an ROI would avoid. Two further technical contributors cannot be excluded: this dataset’s delayed phase at 4–5 min sits at the early end of the equilibrium timing used in the CT-ECV literature, where no consensus exists between roughly 3 and 10 min; and, unlike the aortic mask, tumor masks were not eroded before sampling. Whether a whole-lesion ECV distribution or a representative-ROI value is the more clinically useful quantity is an open question that this tool makes it possible to ask, and that a comparison against matched manual ROI measurement should answer.

### 4.5 Internal consistency check using a hematocrit-independent profile

Because this dataset provides no measured hematocrit, we added a hematocrit-independent check by sampling mean attenuation across all four phases within each tumor. Tumor instances showing a CT washout sign had a lower ECV (median 35.6%) than those without one (median 43.5%).

This relationship must be interpreted with care, and we report it as an internal consistency check rather than as a finding. The two quantities are not statistically independent: both are functions of the tumor’s delayed-phase attenuation, so a tumor that is less dense on the delayed phase will both satisfy the washout criterion and yield a smaller native-to-delayed attenuation change, which is the ECV numerator. Consistent with this, the subgroup difference in ECV is fully accounted for by the subgroup difference in that numerator, with no corresponding difference in the blood-pool denominator (p = 0.25), and the correlation between the continuous washout index and ECV does not survive adjustment for the shared delayed-phase term (partial Spearman ρ = −0.04, p = 0.57). What this check does establish is that the tool’s output responds to delayed-phase attenuation in the direction and magnitude the ECV formula dictates, across an independent 246-instance sample — which is the property one wants to verify in an implementation. It should not be cited as evidence that washout status predicts ECV independently.

### 4.6 Quality control and failure reporting as design features

For a tool intended to run unattended, the informative property is not how often it completes but whether it fails loudly when it should. This evaluation provides three concrete demonstrations. Craniocaudal field-of-view mismatch between phases is pervasive in this dataset, and because the TotalSegmentator aortic label [13] extends into the thoracic aorta well beyond the liver, the blood-pool ROI is considerably more exposed to it than the liver ROI; without the geometry-based slice exclusion of 2.7 these cases returned silently implausible blood-pool values rather than failing, and after the correction they either recover with physiologically sane enhancement or fail with an explicit diagnostic message. Second, and most usefully, the blood-pool validity check identified a mislabeled series in a published, publicly distributed dataset: case 369’s declared unenhanced acquisition is a second reconstruction of the portal venous phase (3.2). A pipeline without that check would have returned a number for this case, and the number would have been meaningless. Detecting a data error is a stronger result for a quality-control design than failing gracefully on a difficult case, and it is the reason we regard the validity thresholds as part of the method rather than as defensive programming. Third, the four retained continuous QC measures (2.8) independently flagged every physiologically impossible value discussed in 4.3 and 4.4, so those cases are identifiable by a downstream user without visual review of all 156.

Two counterweights belong here. The organ-mask lookup bug (3.1) shows that the failure being reported was as often ours as the data’s: an early version of this pipeline reconstructed mask filenames instead of indexing the archive and misreported 29.9% of cases as lacking a segmentation when the true figure was zero. We report it in full because the alternative would have been to publish a limitation of our software as a limitation of a public dataset. And the QC instrumentation is demonstrably not uniformly well calibrated: the aortic attenuation sanity range as implemented (−30 to 150 HU) flags only 3 of 156 cases and did not flag case 369 at 93.9 HU, which is the one case where it would have mattered. That range is better described as a screen for gross error — a mask lying in fat or bone, or a grossly wrong series — than as a physiological range for unenhanced blood, since neither bound corresponds to blood. A check on the fraction of the ROI above a calcium threshold would have separated the two contaminated cases cleanly (12.8% and 35.1% of ROI voxels above 100 HU) and is the more direct measurement of the underlying cause; it is not implemented here. We would argue that a failure-mode taxonomy of the kind in Table 2, including the checks that did not work, belongs in the reporting of any batch imaging pipeline alongside the headline completion rate.

### 4.7 Hematocrit

The absence of a measured hematocrit is inherent to secondary analysis of a public dataset assembled for other purposes, and the application exposes the assumed value as an editable parameter precisely because no single value is defensible for every patient. Substituting a per-patient estimate derived from aortic attenuation moved both liver and tumor ECV further from, not closer to, published ranges (3.7), which we take as indirect evidence against that estimate in this dataset rather than as evidence that a fixed 0.40 is correct for any individual. A related observation bears on the estimator itself: while the modal aortic attenuation is on average more robust to focal plaque than the mean (SD 9.8 vs 24.3 HU), one case returned a modal value of −23 HU, physiologically impossible for blood, which the mean-based sanity range did not catch. Robustness of a summary statistic in aggregate does not guarantee it per case.

### 4.8 Limitations

Registration quality was assessed by qualitative visual review of two representative cases before this study (2.3) rather than by a quantitative metric across the cohort, and the unmodified Elastix “default0” preset [12] was used throughout; every downstream number inherits whatever registration error that preset carries. In-plane field-of-view and slice-thickness mismatch, present in 13% of the full cohort and severe in 3%, is characterized but not corrected, unlike its craniocaudal counterpart; one case with an extreme inter-phase scan-length difference was accepted as an uncorrected limitation. Tumor masks were not eroded before sampling (4.4). Two failures were attributed to aortic plaque or thrombus by targeted visual review rather than by an automated screen. The relationship between the dynamic multiphase pattern and ECV was examined only between ROI-level summaries, not voxel by voxel, although both quantities exist as spatially resolved maps; a per-voxel comparison is the more informative analysis and has not been done. The eight failed cases were not revisited interactively, so whether some could be recovered by manual ROI placement or manual registration adjustment in the application is untested — the reported 95.1% therefore characterizes the unattended batch path, not the limit of what the tool can process. The four-phase dynamic sampling carries no plausibility filter of its own (3.6). The comparisons in Section 3 are descriptive and exploratory, uncorrected for multiple testing, and the reported associations are not independent of the ECV calculation (4.5). Most importantly for a tool report, this is a single-dataset evaluation on retrospective public data: the application has not been evaluated against a reference standard, against matched manual ROI measurement, or for inter-operator reproducibility, and no claim of clinical validity or fitness for diagnostic use is made or implied.

### 4.9 Availability and future work

CT ECV Mapper is not distributed publicly and is not listed in the 3D Slicer Extension Manager. It is made available to research institutions under a collaboration or support agreement; enquiries should be directed to the corresponding author. The dataset analyzed here is publicly available under a CC-BY-4.0 license (doi:10.5281/zenodo.12741586), and the unmodified Elastix parameter files used for every registration reported in this study are archived with the project source so that the registration step can be reproduced independently of the application. Planned work follows directly from the limitations above: a quantitative registration accuracy assessment, an in-plane field-of-view correction to complement the existing craniocaudal one, optional boundary erosion for lesion ROIs, a plausibility filter on the dynamic multiphase sampling, interactive re-examination of the cases the unattended path could not process, and a comparison of whole-lesion ECV distributions against matched manual ROI measurement by readers, which is the natural next step toward establishing what the map adds over the scalar.

### 4.10 Conclusion

CT ECV Mapper makes voxelwise liver and tumor ECV mapping available from conventional single-energy multiphase CT on an open platform, with adjustable parameters and three-dimensional ROI measurement for interactive use and a scriptable path for unattended cohort processing. In an evaluation on 164 patients it completed 95.1% of cases, extended tumor ECV coverage more than sixfold relative to a conventional two-phase approach, produced liver ECV consistent with published values for a cirrhotic population, and failed explicitly and diagnosably in the cases it could not process. The tool is presented as a technically feasible and reportable implementation, not as a validated diagnostic instrument.

## Declarations

## Funding

This work received no public, commercial or institutional grant funding. All costs were borne personally by the author.

## Competing interests

The author is the founder and funder of Suzuki Medical Imaging Lab Co., Ltd. The author developed the software described in this article and intends to make it available to research institutions under a paid support arrangement, as stated in 4.9; this constitutes a financial interest in the software evaluated here. The author is also affiliated with Plusman LLC, which provided no funding, materials or other support for this work.

Ethics approval

The original data collection was approved by the Bioethics Committee at the Medical University of Warsaw (approval AKBE/41/2024). The present study is a secondary analysis of a de-identified dataset released publicly under a CC-BY-4.0 license; no additional institutional review was sought for this use.

## Consent for publication

Not applicable. The dataset contains no identifiable individual data.

Author contributions

M.S. is the sole author and was responsible for the study design, the software specification and development, the data analysis and its interpretation, and the drafting and revision of the manuscript, with the AI assistance described below and in 2.11.

Data and software availability

As stated in 4.9: the dataset analyzed is publicly available (doi:10.5281/zenodo.12741586); the software is not publicly distributed and is available to research institutions under a collaboration or support agreement on enquiry to the corresponding author.

AI use

During the preparation of this work the author used Claude (Anthropic) to assist with drafting and revising the manuscript text, with literature retrieval, and with the software development and analysis described in 2.11. After using this tool the author reviewed and edited all content, verified all reported values against the source data, and takes full responsibility for the content of this article. The tool is not an author and is not cited as one.

## Data Availability

The dataset analyzed is publicly available (doi:10.5281/zenodo.12741586); the software is not publicly distributed and is available to research institutions under a collaboration or support agreement on enquiry to the corresponding author.

https://doi.org/10.5281/zenodo.12741586

## Notes

### Author Declarations

The study used ONLY openly available human data. All data analyzed in this work were obtained from the WAW-TACE dataset, deposited on Zenodo as record 12741586 under a CC-BY-4.0 license and publicly released in 2024, before the initiation of this study. The dataset comprises baseline four-phase abdominal CT of 233 patients with hepatocellular carcinoma treated with transarterial chemoembolization, together with TotalSegmentator-derived organ segmentations, hand-crafted tumor segmentations and clinical metadata, and is described in: Bartnik K, Bartczak T, Krzyzinski M, et al. WAW-TACE: A Hepatocellular Carcinoma Multiphase CT Dataset with Segmentations, Radiomics Features, and Clinical Data. Radiology: Artificial Intelligence 2024;6(6):e240296. The original data collection was approved by the Bioethics Committee at the Medical University of Warsaw (approval AKBE/41/2024) by the group that assembled the dataset. The present work is a secondary analysis of that de-identified public release; no additional data were collected, no identifiable information was accessed, and no attempt was made to re-identify any individual. All files were downloaded directly from the public Zenodo record, with no application, registration, screening or access request of any kind: https://doi.org/10.5281/zenodo.12741586 https://zenodo.org/records/12741586 No other data source was used.

## References

1. Llovet JM, Kelley RK, Villanueva A, Singal AG, Pikarsky E, Roayaie S, Lencioni R, Koike K, Zucman-Rossi J, Finn RS. Hepatocellular carcinoma. Nat Rev Dis Primers. 2021;7:6.

2. Reig M, Forner A, Rimola J, Ferrer-Fàbrega J, Burrel M, Garcia-Criado Á, Kelley RK, Galle PR, Mazzaferro V, Salem R, Sangro B, Singal AG, Vogel A, Fuster J, Ayuso C, Bruix J. BCLC strategy for prognosis prediction and treatment recommendation: the 2022 update. J Hepatol. 2022;76(3):681–693.

3. Chernyak V, Fowler KJ, Kamaya A, Kielar AZ, Elsayes KM, Bashir MR, Kono Y, Do RK, Mitchell DG, Singal AG, Tang A, Sirlin CB. Liver Imaging Reporting and Data System (LI-RADS) Version 2018: imaging of hepatocellular carcinoma in at-risk patients. Radiology. 2018;289(3):816–830.

4. Bandula S, Punwani S, Rosenberg WM, Jalan R, Hall AR, Dhillon A, Moon JC, Taylor SA. Equilibrium contrast-enhanced CT imaging to evaluate hepatic fibrosis: initial validation by comparison with histopathologic sampling. Radiology. 2015;275(1):136–143.

5. Yoon JH, Lee JM, Kim JH, Lee KB, Kim H, Hong SK, Yi NJ, Lee KW, Suh KS. Hepatic fibrosis grading with extracellular volume fraction from iodine mapping in spectral liver CT. Eur J Radiol. 2021;137:109604.

6. Zhang H, Hao E, Xia D, Ma M, Wu J, Liu T, Gao M, Wu X. Estimating liver cirrhosis severity with extracellular volume fraction by spectral CT. Sci Rep. 2025;15:18343.

7. Honda T, Onishi H, Fukui H, Yano K, Kiso K, Nakamoto A, Tsuboyama T, Ota T, Tatsumi M, Tahara S, Kobayashi S, Eguchi H, Tomiyama N. Extracellular volume fraction using contrast-enhanced CT is useful in differentiating intrahepatic cholangiocellular carcinoma from hepatocellular carcinoma. Front Oncol. 2023;13:1214977.

8. He B, Xue L, Wu Z, Guo D, Shen Y, Miao Y. Discrimination of hepatocellular carcinoma from intrahepatic cholangiocarcinoma using extracellular volume fraction (ECV) threshold on contrast-enhanced CT [preprint]. Research Square. Posted 2025-11-11. doi:10.21203/rs.3.rs-7651673/v1. Preprint; not peer-reviewed at the time of writing.

9. Kellman P, Wilson JR, Xue H, Ugander M, Arai AE. Extracellular volume fraction mapping in the myocardium, part 1: evaluation of an automated method. J Cardiovasc Magn Reson. 2012;14:63.

10. Bartnik K, Bartczak T, Krzyziński M, Korzeniowski K, Lamparski K, Węgrzyn P, Lam E, Bartkowiak M, Wróblewski T, Mech K, Januszewicz M, Biecek P. WAW-TACE: a hepatocellular carcinoma multiphase CT dataset with segmentations, radiomics features, and clinical data. Radiol Artif Intell. 2024;6(6):e240296.

11. Fedorov A, Beichel R, Kalpathy-Cramer J, Finet J, Fillion-Robin JC, Pujol S, Bauer C, Jennings D, Fennessy F, Sonka M, Buatti J, Aylward S, Miller JV, Pieper S, Kikinis R. 3D Slicer as an image computing platform for the Quantitative Imaging Network. Magn Reson Imaging. 2012;30(9):1323–1341.

12. Klein S, Staring M, Murphy K, Viergever MA, Pluim JPW. elastix: a toolbox for intensity-based medical image registration. IEEE Trans Med Imaging. 2010;29(1):196–205.

13. Wasserthal J, Breit HC, Meyer MT, Pradella M, Hinck D, Sauter AW, Heye T, Boll DT, Cyriac J, Yang S, Bach M, Segeroth M. TotalSegmentator: robust segmentation of 104 anatomic structures in CT images. Radiol Artif Intell. 2023;5(5):e230024.

14. Kim NY, Im DJ, Youn JC, Hong YJ, Choi BW, Kang SM, Lee HJ. Synthetic extracellular volume fraction derived using virtual unenhanced attenuation of blood on contrast-enhanced cardiac dual-energy CT in nonischemic cardiomyopathy. AJR Am J Roentgenol. 2022;218(3):454–461.

